# Anti-inflammatory effects of 12-HHT via epithelial barrier enhancement in colon organoids of normoganglionosis in Hirschsprung’s disease

**DOI:** 10.64898/2026.02.18.26346528

**Authors:** Kazuto Suda, Kumpei Abe, Yurina Nishimura, Masafumi Tanaka, Yuki Nagasako, Xuxuan Rao, Jianqin Zhang, Siyi Zeng, Kentaro Fujiwara, Shunsuke Yamada, Junya Ishii, Shiho Yoshida, Soichi Shibuya, Go Miyano

## Abstract

**Purpose:** Hirschsprung-associated enterocolitis remains a major postoperative complication of Hirschsprung’s disease (HD), and impaired epithelial barrier integrity has been proposed as a contributing factor. In this study, we investigated whether 12-hydroxyheptadecatrienoic acid (12-HHT), an endogenous leukotriene B4 receptor 2 (BLT-2) agonist, enhances the epithelial barrier and exerts anti-inflammatory effects in patient-derived colonic organoids.

**Methods:** Normoganglionic specimens from rectal/rectosigmoid HD at pull-through (HD-N; n = 8) and transverse colon specimens from anorectal malformation (ARM) at colostomy closure (n = 10) were used to generate colonic organoids. Epithelia were isolated using ethylenediaminetetraacetic acid and subsequently embedded in Matrigel. Baseline expression of *TJP1*, *TJP2*, *F11R* (encoding junctional adhesion molecule-A), *JAM2*, *CLDN1*, *CLDN3*, *CLDN4*) and *LTB4R2* (encoding BLT-2) was assessed by qPCR and immunoblotting. Organoids were then treated with 12-HHT (0.4, 2, or 10 μM) for 7 days, followed by qPCR. Additional experiments assessed cytokine expression (*IL1B*, *IL6*) and TJPs after 24 h with tumor necrosis factor-α (TNF-α, 100 ng/mL) plus phosphate buffered saline or 12-HHT. Barrier function was evaluated using FITC–dextran influx assays.

**Results:** HD-N and ARM organoids exhibited similar growth efficiencies. Baseline expression for *F11R*, *JAM2*, *CLDN1*, *CLDN3*, *CLDN4*, and *LTB4R2* was significantly lower in HD-N than in ARM. TJPs were upregulated by 12-HHT at 2 and 10 μM in both groups, with stronger effects in ARM. In HD-N organoids, 10 μM 12-HHT suppressed TNF-α-induced *IL1B* and *IL6* elevation mitigated tight junction proteins (TJPs) downregulation more effectively than 2 μM. 12-HHT attenuated TNF-α–induced FITC–dextran influx in HD-N organoids.

**Conclusion:** 12-HHT may exert anti-inflammatory effects by integrating TJPs of HD-N.

## Introduction

Hirschsprung’s disease (HD) is a congenital disorder characterized by functional bowel obstruction due to the absence of enteric ganglion cells in the rectum and distal colon [1–4].

The standard surgical treatment is resection of the aganglionic segment followed by pull-through of the proximal normoganglionic bowel [5, 6]. Despite successful surgery, Hirschsprung-associated enterocolitis (HAEC) can occur even in the absence of obstructive symptoms and may be life threatening, with a reported incidence as high as 33% [7]. The risk of HAEC increases with the length of the aganglionic segment, particularly in cases of long segments, total colonic, or extensive aganglionosis [8, 9]. Emerging evidence suggests that immature intestinal barrier function is not limited to the aganglionic segment but may also be present in the normoganglionic colon of patients with HD [10].

We previously reported that the normoganglionic colons of patients with HD showed significantly reduced expression of tight junction proteins (TJPs), particularly Claudin-4, along with decreased levels of its upstream regulator, Leukotriene B4 receptor 2 (BLT-2) [8]. BLT-2 was originally identified as a low-affinity receptor for Leukotriene B4, which enhances epithelial barrier function by facilitating Claudin-4 transcription in several tissues [11]. These alterations suggest that even the ganglionated segment has an intrinsic epithelial barrier vulnerability, which may predispose patients to bacterial translocation and mucosal inflammation. Accordingly, reduced Claudin-4 and BLT-2 expression may be associated with the biological susceptibility of postoperative HAEC.

12-Hydroxyheptadecatrienoic acid (12-HHT), an endogenous ligand of BLT-2 derived from arachidonic acid metabolism, upregulates TJPs such as Claudin-4, and enhances epithelial barrier integrity by promoting the recovery of transepithelial electrical resistance in MDCK II, a kidney epithelial cell line [11]. However, no studies have investigated the effects of 12-HHT on intestinal epithelial barrier function in HD.

Organoid culture systems recapitulate the structural and functional properties of native tissues and have been widely recognized as valuable platforms in regenerative medicine and stem cell biology [12, 13]. Recent reports have utilized gut-derived organoids to evaluate epithelial barrier integrity in HD, offering a promising platform for disease modeling [14, 15]. Based on these insights, we hypothesized that 12-HHT may improve TJPs expression and barrier function in epithelial organoids derived from the normoganglionic colon in patients with HD. This study aimed to investigate the barrier-enhancing and potential anti-inflammatory effects of 12-HHT using patient-derived organoid cultures to propose a novel therapeutic approach to reduce the risk of HAEC following pull-through surgery.

## Materials and methods

### Study design

Patients were recruited between 3 June 2024 and 27 November 2025. The subjects were colonic specimens of the normoganglionic segment from the rectal or rectosigmoid type of HD at pull-through surgery (HD-N) and transverse colon specimens from anorectal malformations (ARM) at colostomy closure, which served as non-HD controls (Fig. 1). Human colonic epithelial organoids were cultured for seven days and subjected to experimental paradigms.

**Fig. 1.**
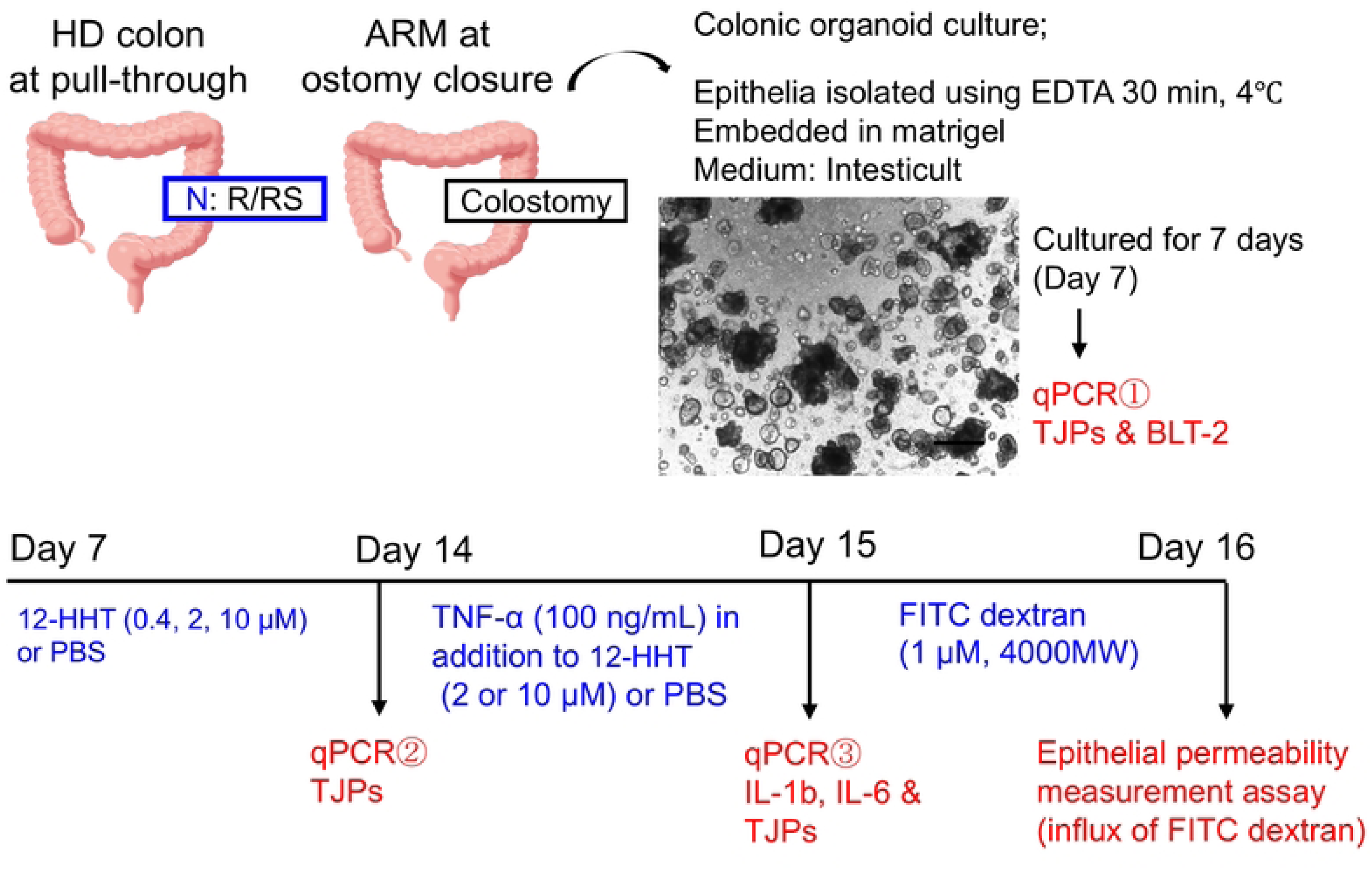
Study design. Epithelial cells were isolated using EDTA treatment from colonic specimens obtained from patients with HD at pull-through surgery and from patients with ARM at the time of ostomy closure. Organoids were cultured for 7 days, followed by baseline qPCR analysis of TJPs and BLT-2. Organoids were passaged and treated with 12-HHT from days 7 to 14, followed by qPCR analysis of the TJPs. Using the same organoid line, organoids were subsequently stimulated with TNF-α in combination with 12-HHT for 24 h, and qPCR for *IL1β*, *IL6*, and TJPs was performed on day 15. Epithelial permeability was evaluated on day 16 using an FITC–dextran influx assay. R/RS; rectal or rectosigmoid colon, N; normoganglionosis.

First, the baseline expression of TJPs and BLT-2 was assessed by qPCR on day 7. In the second paradigm, organoids were passaged and then treated with different concentrations of 12-HHT (0.4, 2, or 10 μM) or phosphate buffered saline (PBS) from day 7 to day 14. TJPs expression was evaluated using qPCR on day 14. Using the same organoid line, organoids were subsequently stimulated with TNF-α (100 ng/mL) in the presence of 12-HHT (2 or 10 μM) or PBS for 24 h, and qPCR for *IL1β*, *IL6*, and TJPs was performed on day 15. Epithelial permeability, assessed in the same organoid line, was measured on day 16 using FITC–dextran influx assay.

### Colonic organoid culture

The human colonic organoids culture was established using a modified protocol based on previously published methods [16, 17]. Briefly, the muscular layer of the resected colon was manually removed using scissors and the remaining tissue was minced and gently rinsed with Dulbecco’s PBS (D-PBS) until the supernatant became clear. After discarding D-PBS, the minced tissue was incubated in 5 mM of Ethylenediaminetetraacetic acid (EDTA) for 30 minutes at 4°C with gentle agitation. Following manual vigorous pipetting, the resulting suspension was passed through a 100-µm cell strainer and centrifuged at 100 × g for 3 minutes at 4°C to isolate epithelial cells. The cell pellet was resuspended and embedded in a 25-µL droplet of Matrigel (Corning, USA), which was placed at the center of each well of a pre-warmed 24-well plate. Organoids were maintained in IntestiCult Organoid Growth Medium (Human) (StemCell Technologies, Canada), which was replaced every three days. For passaging, organoids were harvested from the Matrigel using Cell Recovery Solution (Corning, USA), resuspended in fresh Matrigel, and subsequently replated and cultured in growth medium.

### Reagents treatments (12-HHT and TNF-α)

The BLT-2 agonist 12-HHT (H-1640, Sigma-Aldrich) was prepared as 100 µM aliquots in PBS and used at final working concentrations of 0.4, 2.0, and 10.0 µM in the organoid culture medium. TNF-α was prepared as 10 µg/mL aliquots in PBS, following previously published methods, and applied at a final working concentration of 100 ng/mL [18].

### Quantitative PCR (qPCR)

Total RNA was extracted from the organoid cells using an RNeasy Micro Kit (QIAGEN, Germany). qPCR was performed for TJP-related genes (*TJP1*, *TJP2*, *F11R*, *JAM2*, *CLDN1*, *CLDN3*, *CLDN4*) [19–22], epithelial differentiation and maturation markers, including goblet cell–associated markers (*KLF4*, *TFF3*, and *MUC2*) [23, 24], and pro-inflammatory cytokines (*IL1B* and *IL6*) [25], according to the manufacturer’s specifications. All results were normalized to *GAPDH* conventionally. The primer sequences used for qPCR are listed in Table 1.

**Table 1.**
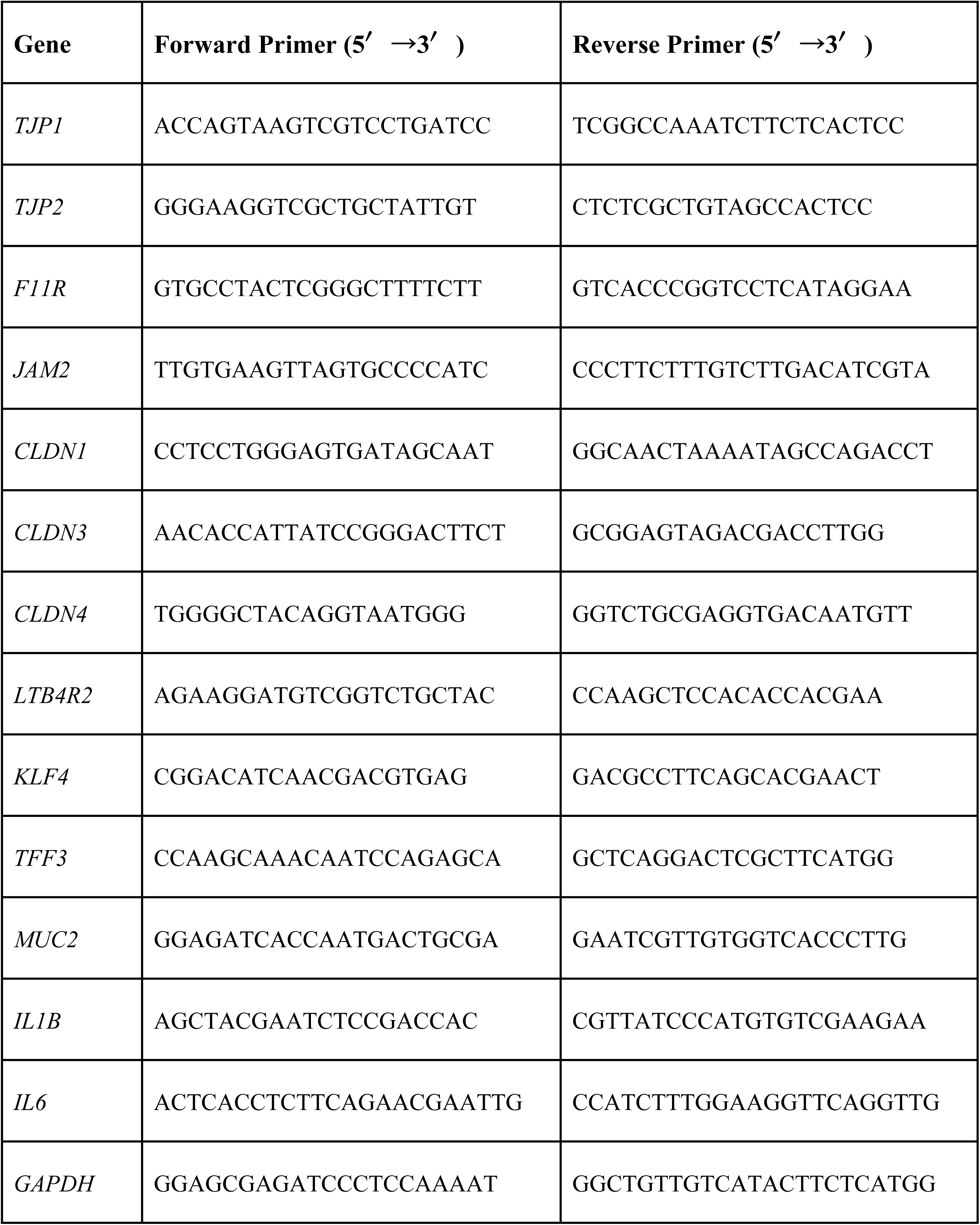
Primer sequences used for qPCR.

### Immunoblotting

Proteins were extracted from organoid cells using RIPA buffer supplemented with a protease inhibitor cocktail (1:50, Roche) to perform immunoblotting according to a previously published technique [8] and the manufacturer’s instructions using the following primary antibodies: CLDN4 (1:5000; Invitrogen) and anti-GAPDH (14C10, 1:5000; Cell Signaling Technology). Signals detected using horseradish peroxidase–conjugated secondary anti-rabbit (1:10,000, Jackson ImmunoResearch) or anti-mouse antibody (1:10,000, Jackson ImmunoResearch) were visualized using Image Quant LAS 4000 (GE Healthcare).

### Quantification of organoid area

For the quantitative analysis of organoid size, five representative images were obtained under each experimental condition. All organoids within each image were analyzed, and the projected area of each organoid was measured using the ImageJ software. The mean organoid area for each condition, including TNF-α treatment with or without 12-HHT, was calculated and normalized to that of the PBS-treated group, which was set to a mean value of 1. Data are presented as relative area ratios.

### Measurement of FITC

FITC–dextran (MW 4000) (MedChemExpress, HY-128868A) was prepared as 1 mM aliquots in dimethyl sulfoxide (DMSO). The permeability of organoid lumens to fluorescent markers was tested using FITC–dextran, according to previously reported methods [26]. Organoids were washed twice with PBS and incubated with FITC–dextran at a final concentration of 1 µM in organoid culture medium for 6 h at room temperature. The marker solution was removed and the organoids were washed five times with PBS. Fluorescence intensity of FITC remaining in the organoid lumen was captured using a BZ-X700 fluorescence microscope (Keyence, Japan) with a GFP filter and quantified using region of interest (ROI) analysis in ImageJ for each image. The mean values were calculated from five images per treatment. Intensities were normalized by subtracting the fluorescence of the PBS + DMSO group (negative control, without TNF-α), and the corrected values were expressed as a ratio relative to the PBS + TNF-α group for graphing.

### Trypan blue assay

Cell viability was assessed using a trypan blue exclusion assay, as previously described [27]. Briefly, cells were trypsinized with TrypLE™ Express Enzyme (Thermo Fisher Scientific, USA; Cat# 12605-010) at 37°C for 5 minutes. The enzymatic reaction was stopped by resuspending the cells in 1 mL of PBS (Nacalai Tesque Inc., Japan; Cat# 27575-31) supplemented with 1% fetal bovine serum (FBS; Thermo Fisher Scientific, USA; Cat# 10270-106). Cells stained with 1 mL of 0.5% trypan blue solution (Nacalai Tesque Inc., Japan; Cat# 29853-34) were immediately loaded onto a hemocytometer (Cat #480200; Corning, USA) and the number of unstained viable cells within a 1 × 1 mm grid was counted under a stereomicroscope.

### Statistics

All data are expressed as mean value ± standard deviations, except for age at specimen collection, which is presented as median (range). Differences between the two groups were tested for statistical significance using an unpaired t-test. All statistical tests were two-sided; *p* value of 0.05 or less was considered statistically significant.

### Ethics

This study was approved by the Institutional Review Board committee of Juntendo University School of Medicine (Institutional Review Board number: E21-0173) and complied with the Helsinki Declaration (2008). Written informed consent was obtained from the parents or legal guardians of all participating minors prior to specimen collection.

## Results

### Patient’s characteristics

Colonic samples were obtained from eight patients with HD and 10 patients with ARM, as summarized in Table 2. The median (range) age at specimen collection differed between groups (HD 2.67 (0.25 - 10.0) years; ARM 0.71 (0.50–1.83) years) (*p* < 0.05). Samples from patients with HD were primarily obtained from the sigmoid colon, whereas samples from patients with ARMs were derived exclusively from the transverse colon.

**Table 2.** Patients’ demographic.

|  | HD-N (n = 8) | ARM (n = 10) | *p* value |
| --- | --- | --- | --- |
| age at specimen collection (year) | 2.67 (0.25 - 10.0) | 0.71 (0.50–1.83) | <0.05 |
| Female:Male | 4:4 | 5:5 | N. S |
| region for sample collection |  |  |  |
| transverse colon | 0 | 10 (100%) | <0.0001 |
| descending colon | 1 (12.5%) | 0 |  |
| sigmoid colon | 7 (87.5%) | 0 |  |

HD-N: normoganglionic specimens from Hirschsprung's disease,
ARM: Anorectal malformation, N.S: not significant.

### Reduced baseline expression of tight junction proteins and BLT-2 in HD-derived organoids

Organoids derived from HD-N and ARM samples exhibited comparable growth efficiencies (Fig. 2A). Baseline analysis of day-7 organoids demonstrated that HD-N-derived organoids showed significantly lower expression of multiple TJP–associated genes than ARM controls, including *F11R*, *JAM2*, *CLDN1*, *CLDN3*, and *CLDN4* (Fig. 2B). Expression of *LTB4R2* (BLT-2) was also markedly low in HD-derived organoids (Fig. 2B). Immunoblotting demonstrated lower Claudin-4 protein levels in HD-N organoids than in ARM organoids (*p* < 0.05) (Fig. 2C, D). Among the differentiation markers, qPCR showed that the expression levels of *TFF3* (*p* < 0.05) and *KLF4* were lower in HD-N organoids than those in ARM organoids, whereas *MUC2* expression was higher in HD-N organoids (*p* < 0.01) (Fig. 2E).

**Fig. 2.**
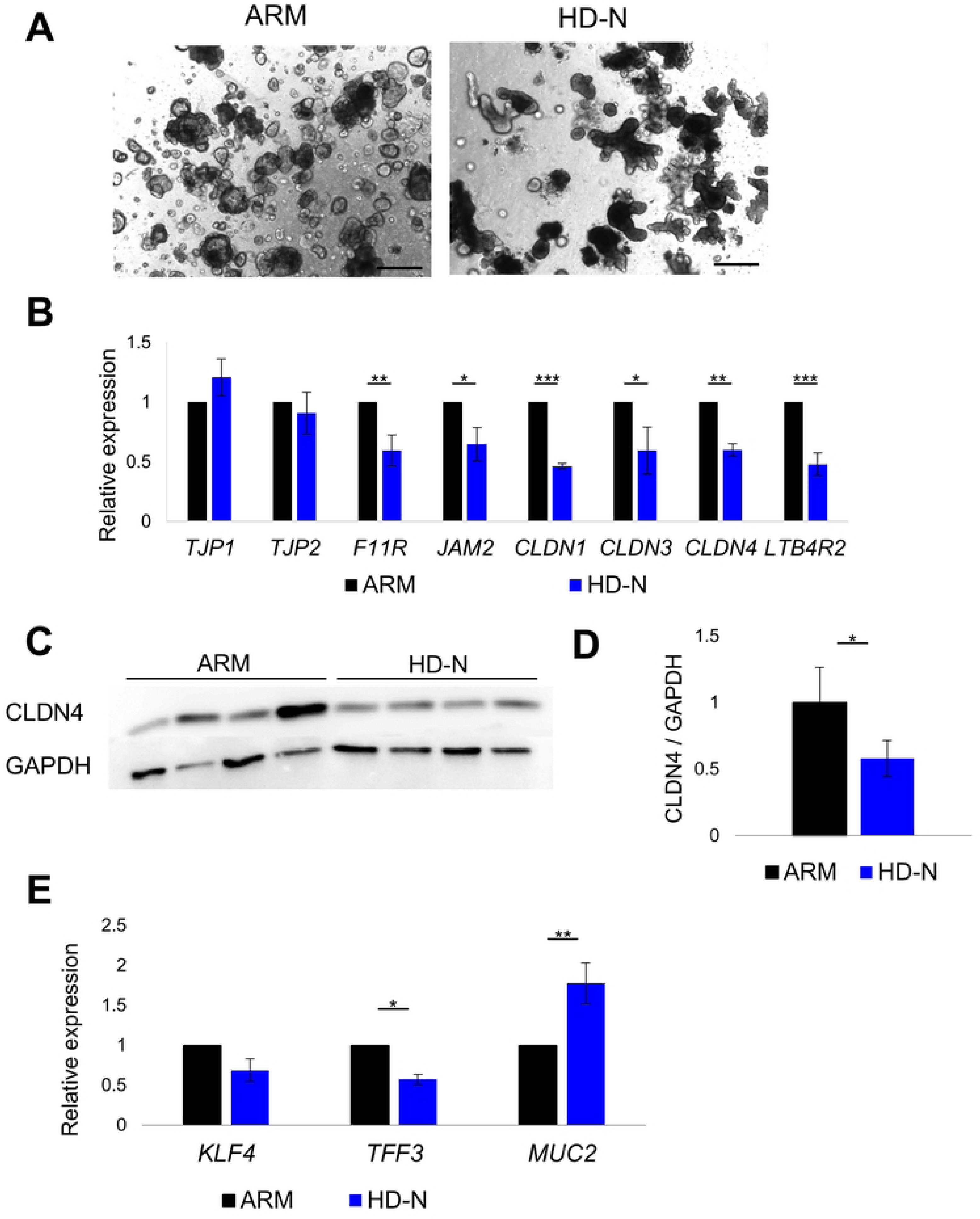
Baseline expression of tight junction–related genes and BLT-2 in HD-N and ARM organoids. A) Representative images of ARM and HD-N organoids formation on days 7. Scale bar: 500 μm. B) Relative expression levels of TJPs and *LTB4R2* (encoding BLT-2) in HD-N and ARM organoids graphed by qPCR. \**p* < 0.05, \*\**p* < 0.01, \*\*\**p* < 0.001. C) Representative immunoblotting images showing Claudin-4 protein levels in ARM and HD-N organoids. D) Quantitative analysis of the CLDN4/GAPDH ratio based on immunoblotting. *p*<0.05. E) Relative expression levels of epithelial differentiation markers in HD-N and ARM organoids graphed by qPCR. \**p* < 0.05, \*\**p* < 0.01.

### 12-HHT modulates not only TJP expression but also differentiation on colonic organoids culture

Treatment with the BLT-2 agonist 12-HHT from day 7 to day 14 resulted in a dose-dependent upregulation of several TJP genes in HD-N organoids, with significant increases observed particularly for *CLDN3* and *CLDN4* at 2 µM and 10 µM (Fig. 3A). This upregulation was more pronounced in ARM organoids (Fig. 3B). No significant morphological changes were observed in the HD-N organoids after 12-HHT treatment (Fig. 3C). The mRNA expression levels of *TFF3*, *KLF4*, and *MUC2* were upregulated by 12-HHT treatment in both the ARM and HD-N groups, with a more pronounced increase observed in the ARM group (Fig. 3D, E).

**Fig. 3.**
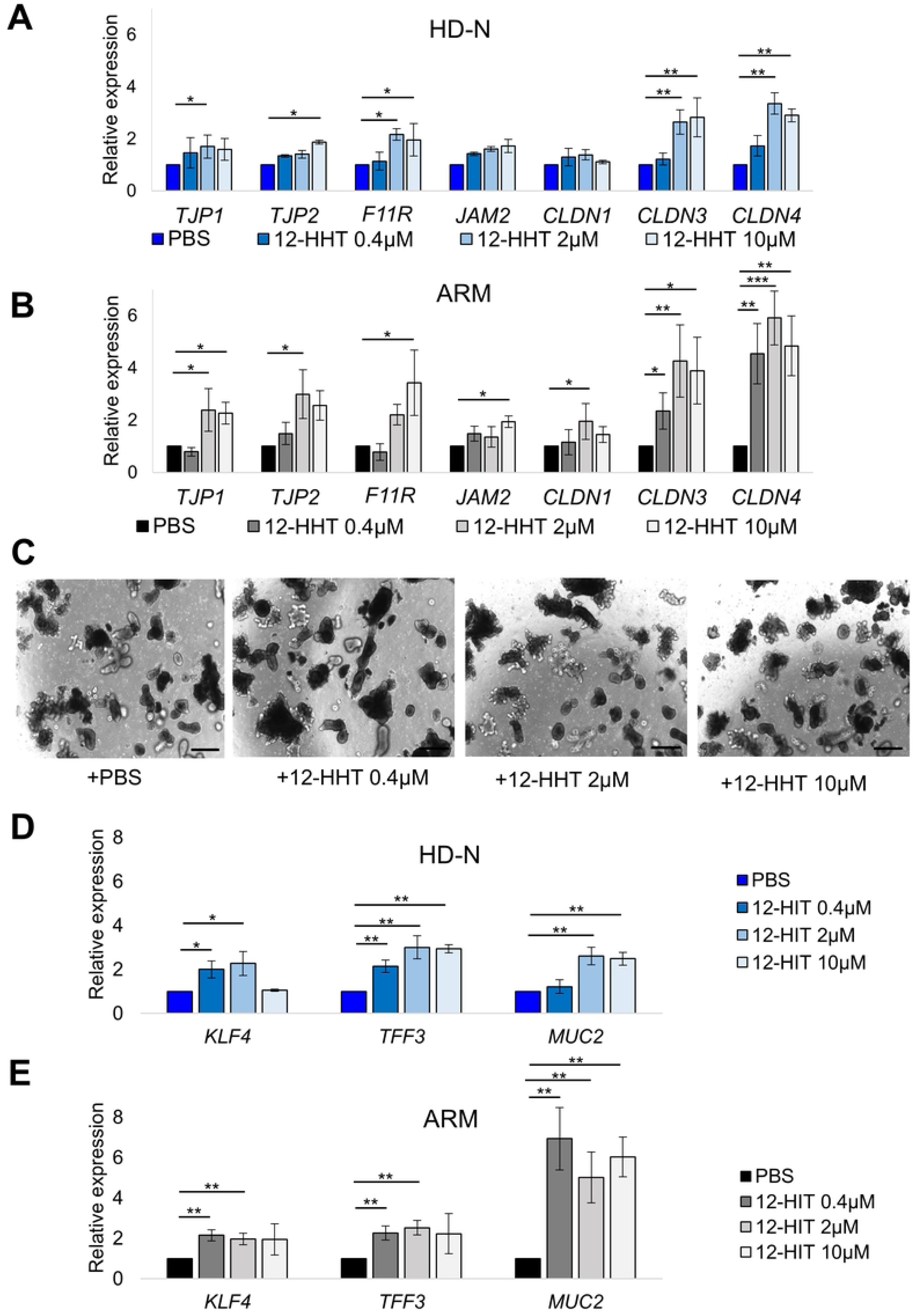
Effects of 12-HHT on barrier-related gene expression in HD-N and ARM organoids. A) Dose-dependent changes in TJP gene expression in ARM organoids treated with 12-HHT (0.4, 2, or 10 µM) for 7 days (day 7–14). \**p* < 0.05, \*\**p* < 0.01. B) Dose-dependent changes in TJP gene expression in HD-N organoids treated with 12-HHT (0.4, 2, or 10 µM) for 7 days (day 7–14). \**p* < 0.05, \*\**p* < 0.01, \*\*\**p* < 0.001. C) Organoids morphology treated with 12-HHT (0.4, 2, or 10 µM) for 7 days (day 7–14). Scale bar: 500 μm. D) Expression of epithelial differentiation markers in HD-N organoids treated with 12-HHT (0.4, 2, or 10 µM) for 7 days (day 7–14). \**p* < 0.05, \*\**p* < 0.01. E) Expression of epithelial differentiation markers in ARM organoids treated with 12-HHT (0.4, 2, or 10 µM) for 7 days (day 7–14). \*\**p* < 0.01.

### The anti-inflammatory effects of 12-HHT on colonic organoids culture

Exposure of organoids to TNF-α for 24 h induced robust upregulation of inflammatory cytokines *IL1β* and *IL6* and reduced expression of multiple TJP genes (Fig. 4A, B). Co-treatment with 12-HHT significantly suppressed TNF-α–induced cytokine expression (Fig. 4A) and restored TJP expression in a dose-dependent manner (Fig. 4B). Representative morphological images and quantitative analysis showed that TNF-α treatment, reduced both the number and mean area of organoids, whereas 12-HHT treatment ameliorated these reductions (Fig. 4C, D). Consistent with the morphological data, trypan blue assays demonstrated that TNF-α stimulation decreased viable cell counts, an effect that was reversed by co-treatment with 12-HHT (Fig. 4E).

**Fig. 4.**
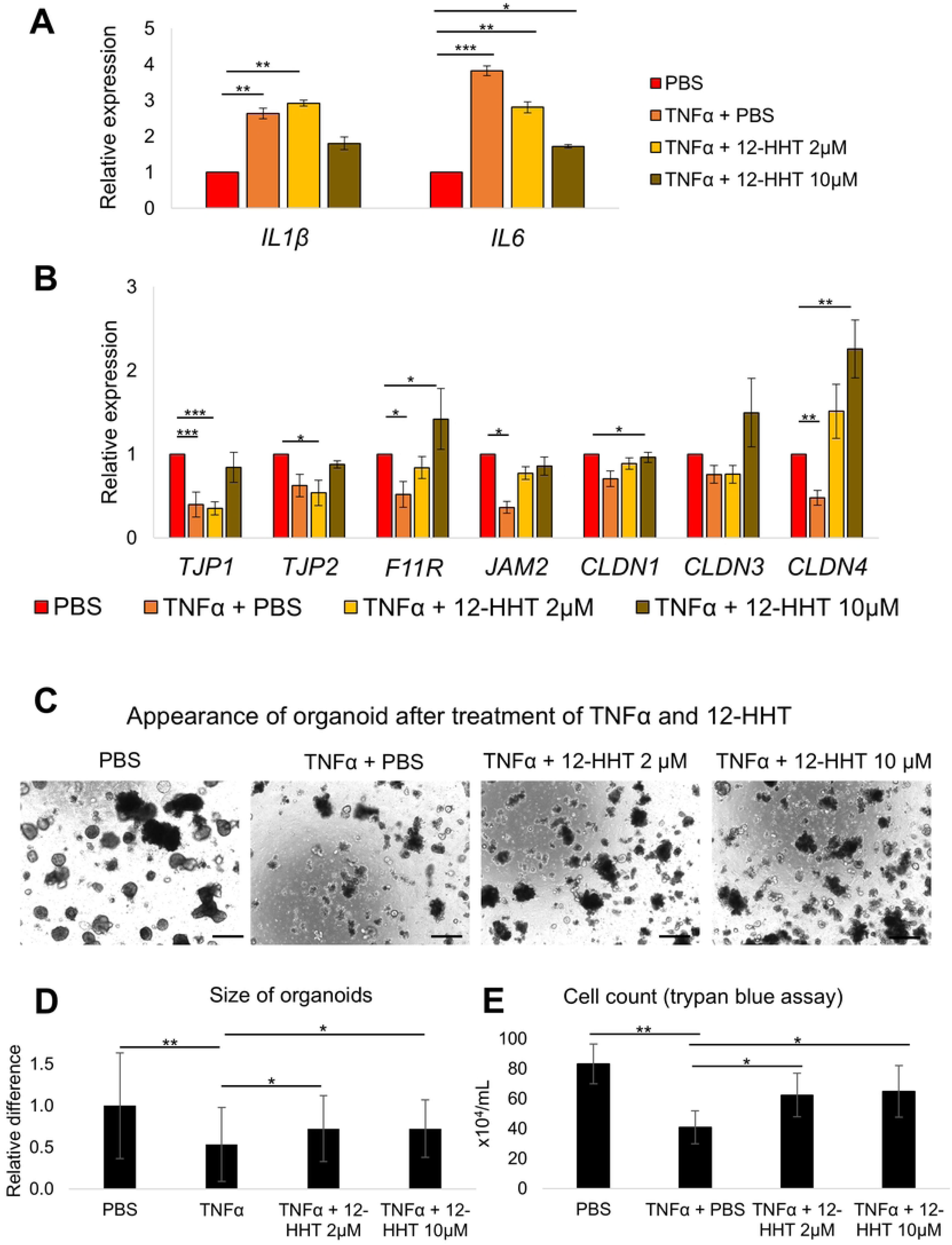
Protective effects of 12-HHT against TNF-α–induced inflammatory signaling and barrier dysfunction in HD-N organoids. A) Relative expression of inflammatory cytokines after 24-h stimulation with TNF-α (100 ng/mL) with or without 12-HHT (2 or 10 µM). * *p*< 0.05, \*\**p* < 0.01, \*\*\**p* < 0.001. B) Relative expression of tight junction genes after 24-h stimulation with TNF-α (100 ng/mL) with or without 12-HHT (2 or 10 µM). \**p* < 0.05, \*\**p* < 0.01, \*\*\**p* < 0.001. C) Representative images of organoid morphology after 24-h stimulation with TNF-α (100 ng/mL) with or without 12-HHT (2 or 10 µM). Scale bar: 500 μm. D) Quantitative analysis of organoid size based on ImageJ measurements. \**p* < 0.05, \*\**p* < 0.01. E) Quantitative analysis of cell counts determined by the trypan blue exclusion assay. \**p* < 0.05, \*\**p* < 0.01.

### 12-HHT improves permeability of colonic organoids

FITC–dextran influx assays demonstrated markedly increased epithelial permeability in HD-derived organoids by TNF-α (Fig. 5A). Co-treatment with 12-HHT significantly reduced FITC influx, with the strongest effect observed at 10 µM (Fig. 5A, B). These findings indicate that BLT-2 activation mitigates TNF-α–induced impairment of epithelial barrier.

**Fig. 5.**
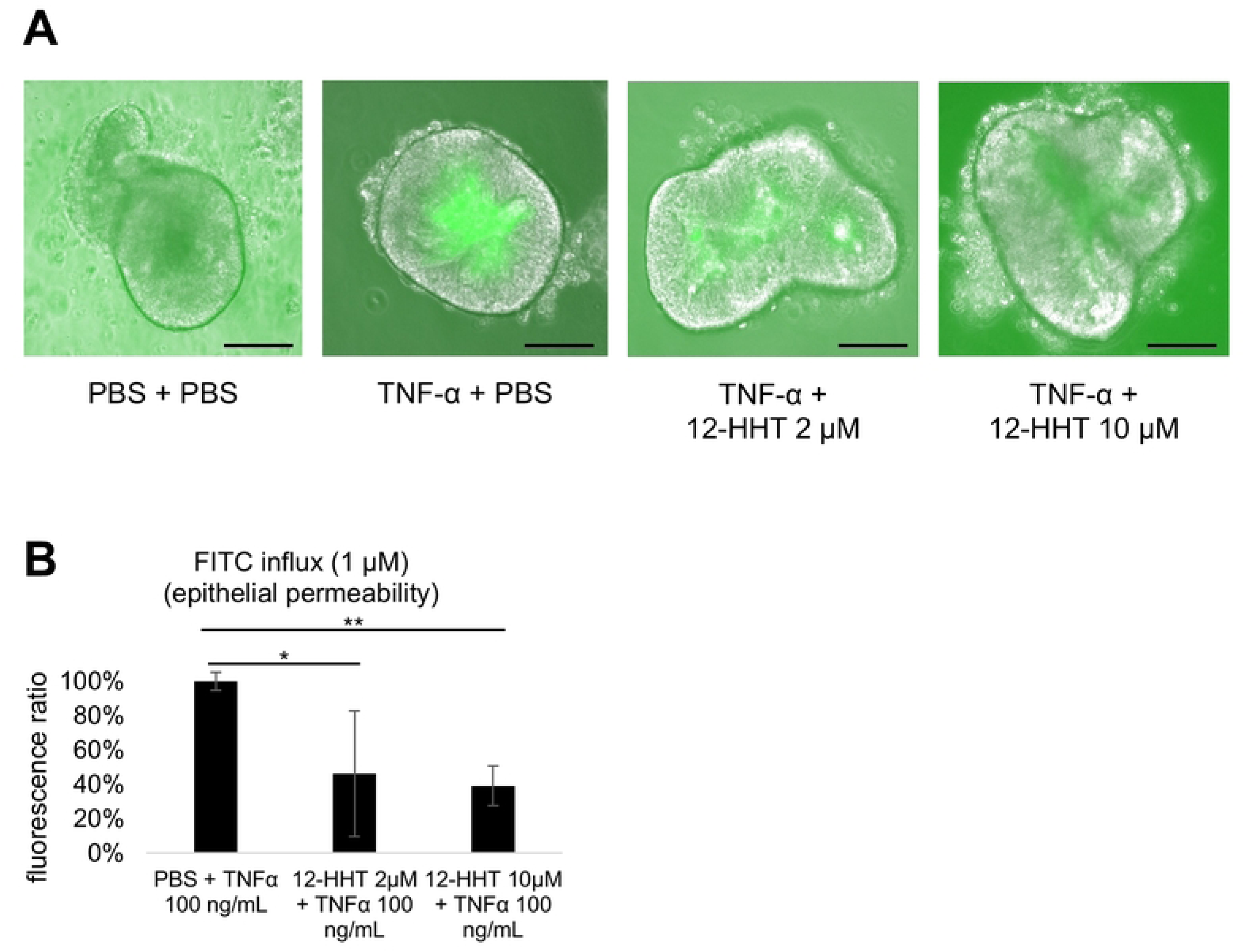
Reduction of TNF-α–induced epithelial permeability by 12-HHT in HD-N organoids. (A) Representative fluorescence images of FITC–dextran (1 µM) influx after 24-h exposure to TNF-α with or without 12-HHT in HD-N organoids. Scale bar: 100 μm. (B) Quantitative analysis of FITC–dextran influx in HD-N organoid based on ImageJ measurements. \**p* < 0.05, \*\**p* < 0.01.

## Discussion

This study demonstrated that colonic organoids from the normoganglionic segment of patients with HD exhibit intrinsic epithelial barrier impairment, characterized by reduced expression of multiple TJPs and their upstream regulator, BLT-2. Importantly, the activation of BLT-2 by its endogenous ligand 12-HHT upregulated TJPs expression, suppressed pro-inflammatory cytokine induction, and retained epithelial permeability even under inflammatory stress. These findings extend our previous observations in patient tissues and provide functional evidence that epithelial vulnerability in HD is not restricted to the aganglionic segment, but persists in the normoganglionic bowel [8]. Collectively, our results suggest that impaired BLT-2–mediated barrier regulation may contribute to HAEC susceptibility and pharmacological reinforcement of epithelial integrity may offer a promising therapeutic concept. The novelty of the present study lies in demonstrating that epithelial barrier dysfunction in the normoganglionic colon can be functionally restored under inflammatory stress through BLT-2 activation, using human gut-derived organoids from patients with HD. Through the introduction of an organoid culture system, we experimentally validated this concept in a human context, which could not be directly examined in vivo.

Interestingly, the TJP-activating effect of 12-HHT was more pronounced in the ARM organoids than in HD-N. This is likely attributable to the low baseline expression of BLT-2 in the HD-N organoids, which may allow for less efficient activation of TJPs. These findings suggest that the magnitude of the 12-HHT effect depends on the baseline level of BLT-2 expression. Similarly, the activation of the differentiation markers *KLF4*, *TFF3*, and *MUC2* in response to 12-HHT was less pronounced in HD-N organoids than in ARM organoids. This attenuated response may be attributable to disease-specific epithelial vulnerability in the normoganglionic colon of patients with HD, whereas differences in baseline BLT-2 expression levels may also contribute to the observed effects.

Previous studies reported findings consistent with our observations regarding the effects of BLT-2 activation mediated by 12-HHT. In a dextran sodium sulfate–induced murine colitis model, BLT-2-deficient mice exhibited more severe disease, characterized by greater body weight loss, enhanced intestinal inflammation, increased expression of pro-inflammatory cytokines, and augmented accumulation of activated macrophages [28]. Moreover, the 12-HHT–BLT-2 axis enhances epithelial barrier function by upregulating Claudin-4 expression and promoting the recovery of transepithelial electrical resistance by activating the p38 MAPK pathway or the p38–PKC signaling axis [11, 29]. Collectively, these studies support the protective role of BLT-2 against inflammatory injury, whereby BLT-2 signaling strengthens the integrity of the epithelial barrier in the intestine and other epithelial tissues, thereby conferring resistance to inflammatory insults. Furthermore, KLF4, a transcription factor that promotes the terminal differentiation of intestinal epithelial cells; TFF3, a goblet cell–derived secretory peptide involved in mucosal protection and epithelial restitution; and MUC2, a mucin produced by mature goblet cells [23, 24], were transcriptionally activated by 12-HHT in both HD-N and ARM organoids. Previous studies have demonstrated that 12-HHT signaling promotes epithelial repair and cell migration [30, 31]. However, its direct role in the regulation of epithelial differentiation and maturation remains unclear. Indeed, even in our experimental system, it is difficult to determine whether 12-HHT directly promotes epithelial differentiation, or whether its epithelial protective effects preserve differentiated cell populations, thereby resulting in the increased expression of differentiation-associated markers. Additionally, the baseline expression levels of BLT-2 and Claudin-4 were lower in the HD-N organoids than in the ARM organoids. A subclinical inflammatory state has been reported in the intestinal epithelium of patients with HD, even without active enterocolitis [32, 33]. Thus, reduced expression of differentiation markers such as *KLF4* and *TFF3* in HD-N organoids may reflect an underlying epithelial vulnerability associated with chronic, low-grade inflammatory stress rather than an active inflammation.

This study has some limitations. First, patients were not stratified by postoperative HAEC history because the cohort predominantly included short-segment HD cases, which have a relatively low HAEC incidence; none developed HAEC before pull-through surgery. Second, obtaining colonic specimens from healthy children is ethically challenging. In this context, ARM tissue has been widely used as a clinically realistic control because it is associated with fewer inflammatory background abnormalities than HD [8, 33, 34]. Although the anatomical location and age at specimen collection differed between the HD and ARM groups in the present study, previous reports, including ours, have provided limited evidence that these factors alone account for the marked differences observed in the expression of tight junction proteins such as Claudin-4 [8, 35, 36]. Moreover, the organoid model represents epithelial cells alone and does not incorporate immune cells, neuronal components, or intestinal microbiota, all of which are recognized modulators of HAEC pathophysiology. Accordingly, more complex systems, such as co-culture approaches or microfluidic organ-on-chip models, may be required to better elucidate the interactions among epithelial cells, immune cells, and microbiota in HD. In addition, the relatively weaker response to 12-HHT observed in HD-N organoids compared to ARM organoids warrant further investigation, including evaluation of higher ligand concentrations, longer treatment durations, and alternative strategies for BLT-2 activation.

In conclusion, our findings suggest that BLT-2 activation contributes to the restoration of epithelial barrier integrity in HD-derived tissues. Enhancing epithelial resilience may represent a novel therapeutic strategy to reduce the susceptibility to HAEC after pull-through surgery, particularly in patients with sustained impairment of epithelial integrity.

## Data Availability

All relevant data are within the manuscript and its Supporting Information files.

## Acknowledgments

The authors thank the Laboratory of Molecular and Biochemical Research, Research Support Center, Juntendo University Graduate School of Medicine, for technical assistance and Maika Ozawa for assistance with the experiments. This study was supported by the Japan Society for the Promotion of Science KAKENHI (24K11813), a Research Grant from the Kobayashi Foundation, and Kawano Masanori Memorial Public Interest Incorporated Foundation for Promotion of Pediatrics Grant Number (33-waka21). We would like to thank Editage (www.editage.jp) for English language editing.

## Author contributions

KS and KA designed this study. SS and GM provided conceptual advice. KA, MT, SY, JI, SY, and SS collected clinical data and samples. KS, KA, YT, YN, XR, and JZ performed the experiments and analyzed the data. KS and KA wrote the manuscript. All the authors have read and approved the final version of the manuscript.

